# Electronic health record mining reveals effects of patient immune status on clinical outcomes of biomaterial implantation following skeletal muscle damage

**DOI:** 10.1101/2025.09.16.25335883

**Authors:** Sonakshi Sharma, Isabella Horton, Aditya Josyula, Kaitlyn Sadtler

**Affiliations:** Section on Immunoengineering, National Institute for Biomedical Imaging and Bioengineering, National Institutes of Health, Bethesda MD 20894; College of Medicine, Pennsylvania State University, Hershey, PA17033; Fischell Department of Bioengineering, University of Maryland, College Park MD 20742; School of Medicine, University of Maryland, Baltimore 21201

## Abstract

Implantation of medical devices and biomaterials can help restore form and function of missing or damaged tissue. It is known that the immune system plays a critical role in both positive and negative outcomes of these implanted materials. The foreign body response is characterized by protein deposition and clotting followed by macrophage inflammation, frustrated phagocytosis, giant cell formation, and ultimately fibrosis. This can inhibit the function of implanted devices (e.g. Insulin pumps) as well as cosmesis (e.g. capsular contracture in breast implants) and persistent inflammation has been associated with more severe outcomes in some patients, including emergence of autoimmune-like pathologies. On the other hand, the immune system plays a constructive role in tissue remodeling and regeneration and is needed for the positive effects of some biomaterials, such as extracellular matrix-based scaffolds in muscle repair. Given these factors, we sought to understand potential variations in post-operative complications in individuals with primary and secondary immune disorders – both autoimmune and immunodeficiencies. This preliminary observational study using electronic health record mining showed increased complication odds for individuals with both autoimmune conditions and immunodeficiencies, with variations dependent upon the individual’s sex and age as well as the type of material implanted. Future prospective studies could yield improved insight into both mechanisms of immune response to materials in humans and identify potential risk factors for individual patients undergoing plastics and reconstructive surgeries.

## INTRODUCTION

Soft tissue damage occurs both due to traumatic injuries and surgical damage. In the United States alone, there are roughly 43.5 million emergency department visits for injuries per year (National Hospital Ambulatory Medical Care Survey: 2022 Emergency Department Summary Tables, CDC (1)) and 51.4 million surgical procedures(National Hospital Discharge Survey 2019, CDC (2)). This damage can result in scarring and permanent defects including volumetric muscle loss (VML) in which 20% or more of the muscle volume is lost resulting in damage beyond the muscle’s regenerative capacity (3). Surgical VML includes muscle resection due to infection, malignancy, or for reconstructive purposes such as muscle flap harvest. Traumatic VML, in the United States, most commonly occurs due to automobile accidents. Regardless of mechanism, the rapid and extensive damage sustained in VML results in inflammation, scarring, and loss of functional capacity of the affected tissues. No effective treatments for extensive muscle loss currently exist, and, as such, the health burden of VML is cumulatively quite significant(4). Recently explored treatment strategies for VML include implantation of various biomaterials to enhance muscle regeneration, including decellularized extracellular matrix (ECM) scaffolds, mesenchymal stem cell scaffolds and more (5). While some of these materials have shown promising results, none have been successful at fully restoring baseline functionality. Dysfunctional wound healing and local immune responses following VML have thus been an area of interest in this field.

During muscle regeneration and healing, there is a crucial transition from a pro-inflammatory to a pro-regenerative immune environment. Dysfunction in this process can result in aberrant wound healing with excessive fat deposition and scar tissue formation. Tissue engineering approaches aim to modulate this response to enhance pro-regenerative immune processes(6). For example, ECM scaffolds are known to modulate the innate immune response to promote tissue repair. In immunologically intact animal models of VML, these scaffolds can reduce fibrosis and enhance constructive inflammation (e.g. M2 macrophage polarization), facilitating muscle fiber regrowth and functional recovery(5, 7–11). Other biomaterials, such as synthetic hydrogels can also promote regeneration given their properties of tunable porosity and biocompatibility which allow these materials to enhance immune cell infiltration and minimize foreign body responses, respectively. In animal models, microporous hydrogels elicited negligible fibrotic encapsulation and instead promoted regenerative macrophage polarization, angiogenesis, and even innervated myofiber formation within the scaffold. On the other hand, non-degradable materials like silicone elicit a strong foreign body response, leading to persistent low-grade inflammation which, over time, causes fibrous capsule formation(12–14).

The host’s response to any of these biomaterials is affected by immune status. Immune status (immunodeficient, immunocompetent, or autoimmune) has been shown to impact biomaterial-facilitated wound healing and muscle repair in preclinical models. A study using tissue-engineered muscle grafts demonstrated superior neovascularization, scaffold integration, and myogenesis in immunocompetent mice compared to immunocompromised mice, underscoring the importance of an intact immune system for muscle repair(6). Conversely, an autoimmune or hyper-inflammatory immune phenotype can skew healing toward maladaptive outcomes. In an autoimmune-prone mouse model, VML injuries treated with biologic ECM scaffolds exhibited attenuated anti-inflammatory (M2) macrophage polarization and instead showed excessive intramuscular fat deposition at the site of injury, indicating aberrant remodeling(8).

Despite these advances in knowledge, there is a notable lack of studies that directly compare the clinical outcomes of different biomaterials used in the presence of damaged skeletal muscle tissue across varying immune statuses. Specifically, empirical data assessing how patients with autoimmune or immunodeficient conditions respond to treatments with materials like ECM scaffolds, hydrogels, or silicone implants is limited. A clearer understanding of how these materials perform in the contexts of immune suppression or autoimmunity is needed to tailor clinically applicable regenerative therapies. As such, this study aimed to evaluate the efficacy of specific biomaterials in skeletal muscle reconstruction across different immune phenotypes.

## METHODS

### Study Design and Population

We conducted a retrospective cohort study of patients who had a history of significant muscle injury due to a variety of mechanisms. Patients whose injuries were treated with specific biomaterials were selected. The data used in this study was accessed in March 2025 from the TriNetX Global Network, which provides access to electronic medical records (diagnoses, procedures, medications, laboratory values, genomic information) from approximately 150 million patients from 149 healthcare organizations worldwide. Patients who met index event criteria (volumetric muscle loss) greater than 20 years ago were excluded. Past medical history data collected included history of chronic kidney disease (CKD), type 2 diabetes mellitus (T2DM), smoking, overweight/obesity, chronic viral infections (e.g., Hepatitis C), and malignancy. Patients were also stratified into age categories: 18-35, 35-50, 50-65, and 65+ years of age for analysis of age-based variations.

### Exposure Definitions and Treatment Groups

Individuals were included if they were older than 18 years of age and had skeletal muscle damage, identified using ICD-10 and CPT codes (**Figure 1**). Skeletal muscle damage codes included surgical or traumatic injury involving extremities, chest wall, and abdominal wall. Adult muscle damage patients were then classified into cohorts based upon the type of biomaterial they were treated with: extracellular matrix-based scaffolds, hydrogels, and silicone-based implants. Within each biomaterial cohort, patients were further subclassified by immune status: autoimmune, immune deficient, and immune competent. Immunodeficiency was defined as a diagnosis of either primary (e.g., common variable immunodeficiency) or secondary immunodeficiency (e.g., HIV, drug or chemotherapy-related immune suppression). Autoimmune conditions included diagnoses such as systemic lupus erythematosus, rheumatoid arthritis, inflammatory bowel diseases, and more. Immunocompetent controls, with immunocompetency defined as not having autoimmune or immune deficient diagnoses, were selected for baseline comparisons. Muscle damage mechanisms were stratified into the following groups: debridement, flap harvest, traumatic injury, and surgical resection. Post-operative outcomes were then compared overall and by immune subgroup to evaluate incidence rates of complications.

**Figure 1.**
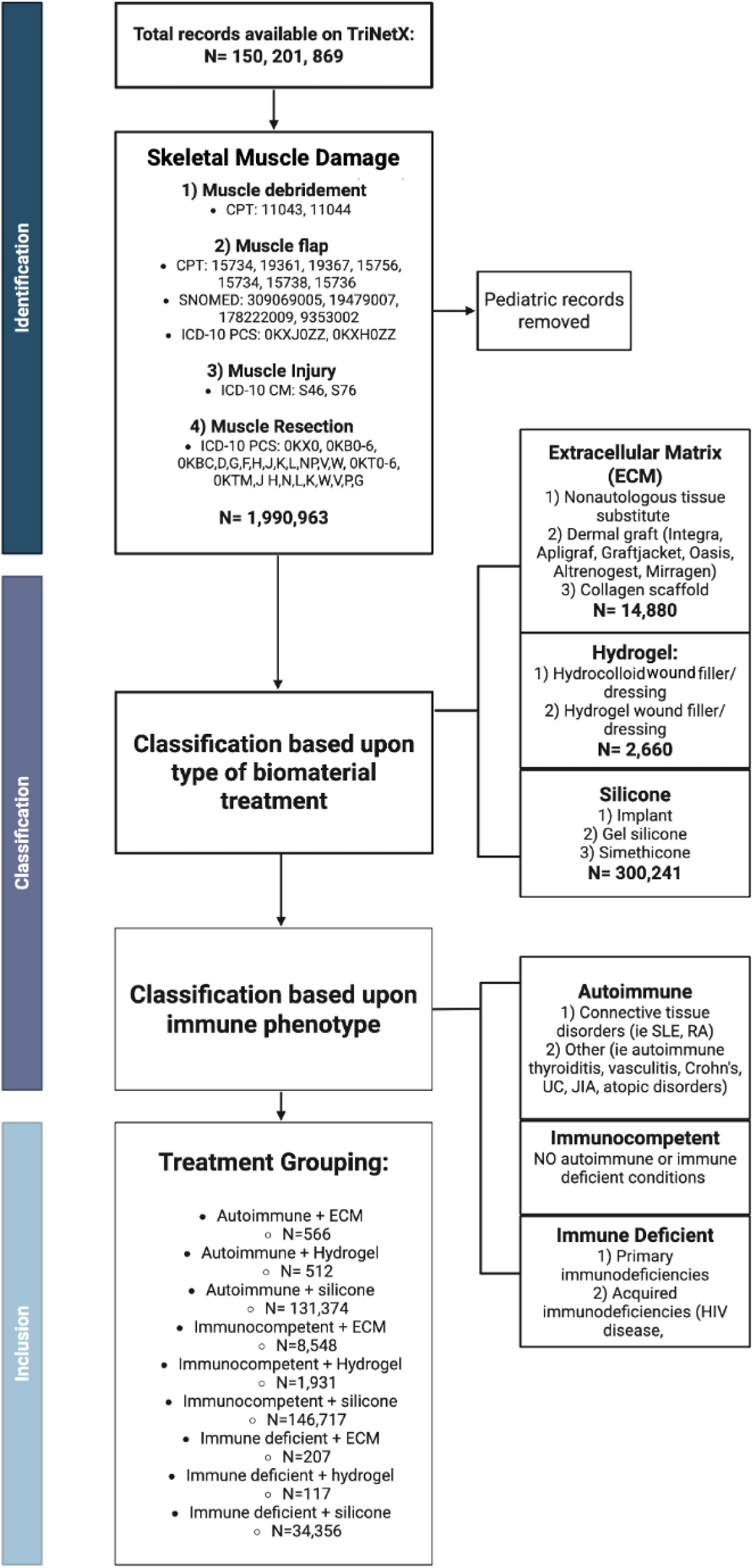
Study design and cohort characteristics.

### Outcome Measures

Primary post-operative outcomes assessed included local tissue edema, post-operative fever, sepsis, elevation of erythrocyte sedimentation rate (ESR) or c-reactive protein (CRP), wound dehiscence, post-operative infection, tissue ischemia, hematoma and seroma formation, cellulitis/ lymphadenitis, mortality and overall graft complication rate. Outcome data were extracted from electronic health records using CPT/ ICD-10 diagnostic codes (**Table 1**). Secondary measures collected included demographic information and past medical history factors relevant to wound healing (e.g. medications, cancer history, smoking history, diabetes, obesity, etc.).

**Table 1.** ICD-10 Codes selected for complications.

| Outcome Variable | Code Source | Codes Included |
| --- | --- | --- |
| Local Tissue Edema | ICD-10 | R60.0 |
| Post-operative Fever | ICD-10 | R50.82 |
| Sepsis | ICD-10 | R65.2 |
| ESR/CRP Elevation | ICD-10 | R79.82, R70 |
| Wound Dehiscence | ICD-10 | T81.3 |
| Post-Operative Infection | ICD-10 | T81.4 |
| Post-Operative Bleeding | ICD-10 | R58, T84.83, T81.7 |
| Post-Operative Ischemia | ICD-10 | R23.1, T79.6 |
| Mortality | ICD-10 | R99 |
| Post-Operative Hematoma | ICD-10 | L76.32 |
| Post-Operative Seroma | ICD-10 | L76.34 |
| Cellulitis/Lymphadenitis | ICD-10 | R59, R22, L03, L04 |
| Graft Complications | ICD-10 | T84, T85 |

### Data and Statistical Analysis

The incidence of each outcome was compared between immune subgroup for all biomaterial types. Secondary analyses included comparison of outcomes based upon sex differences, age differences, menopausal status, type of immune deficiency, and type of volumetric muscle loss sustained. Unadjusted odds ratios (ORs), risk ratios (RR) and corresponding 95% confidence intervals (CIs) were calculated for each outcome and comparison through the TriNetX Global Network platform. Analyses were stratified by biomaterials and immune phenotypes. Forest plots were generated to visualize the relative risk of complications across groups. Zero values were excluded from figures to accommodate the log-transformed scale required for plotting. Statistical analysis was performed using R version 4.5.0. Generation of p values and determination of significance was limited given the number of co-variates and exploratory/observational nature of the study.

## RESULTS

### Demographics by biomaterial group

Demographic data was collected for a total of 24,929 patients (**Table 2**). Mean ages of the groups receiving different biomaterials ranged from 53-63 years of age. While sex distribution was approximately even, the silicone-treated groups had a predominance of females, likely due to use of silicone implants in breast reconstruction. Overall, the patient cohorts were predominantly White and non-Hispanic/Latino. Smoking history, malignancy history and history of chronic viral infection was similar across groups (**Table 3**). History of obesity (range 8%-32%), CKD (range 8%-36%), and T2DM (range 15%-52%) varied widely. Silicone treated patients had a pattern of lower comorbidities while ECM treated patients had higher rates of comorbid conditions.

**Table 2.**
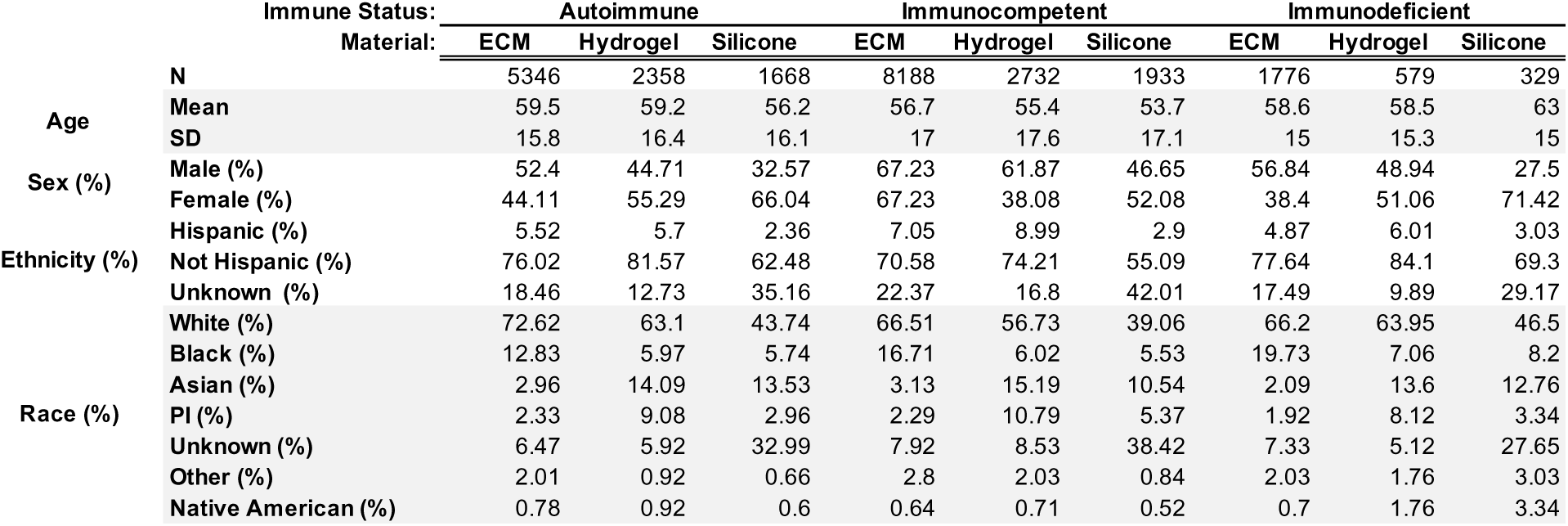
Patient Demographics.

**Table 3.**
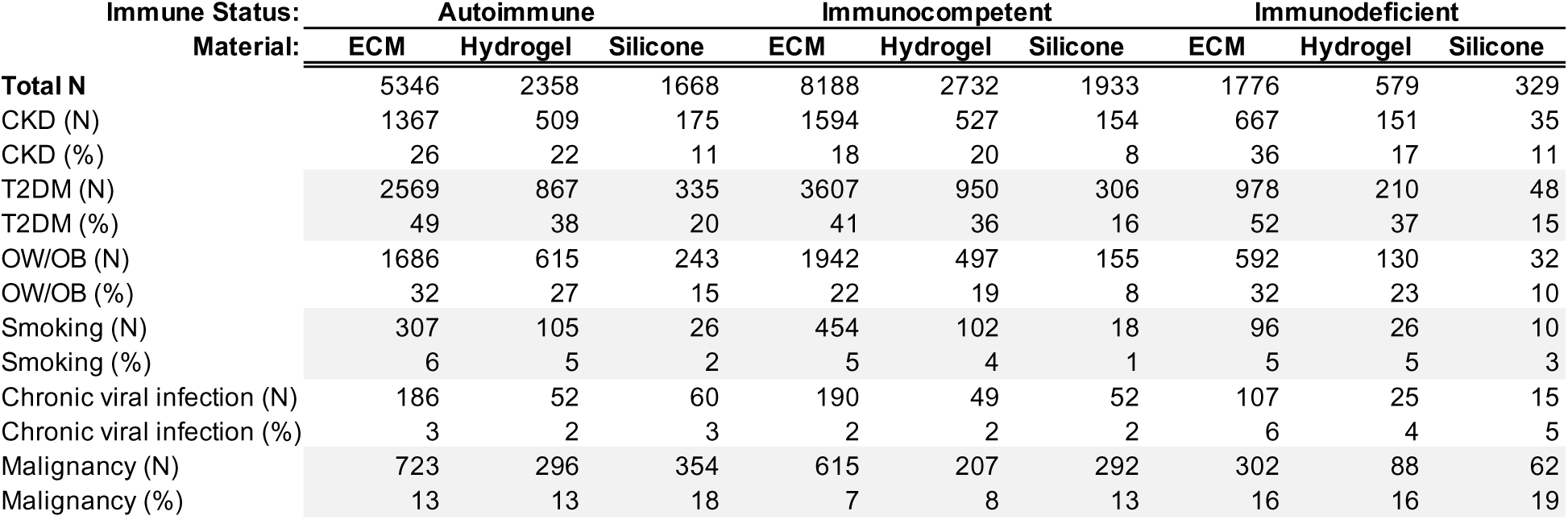
Patient Comorbidities.

### Patient immune status is associated with post-operative complications

In univariable analysis, individuals with autoimmune and immunodeficiencies treated with ECM had higher odds of developing local tissue edema, sepsis, ESR/CRP elevation, wound dehiscence, post-operative infection, post-operative bleeding, tissue ischemia cellulitis/ lymphadenitis, and overall graft complications compared to immunocompetent controls (**Figure 2**). However, the odds of developing complications among immune subgroups did not differ from each other apart from sepsis, which showed higher odds for the immunodeficient group compared to the autoimmune group. Additionally, the autoimmune cohort had higher odds of developing post-operative fever (OR= 1.948, 95% CI, 1.162-3.261) and seroma (OR= 1.762, 95%CI, 1.05-2.954) compared to immunocompetent controls while the immunodeficient group did not.

**Figure 2.**
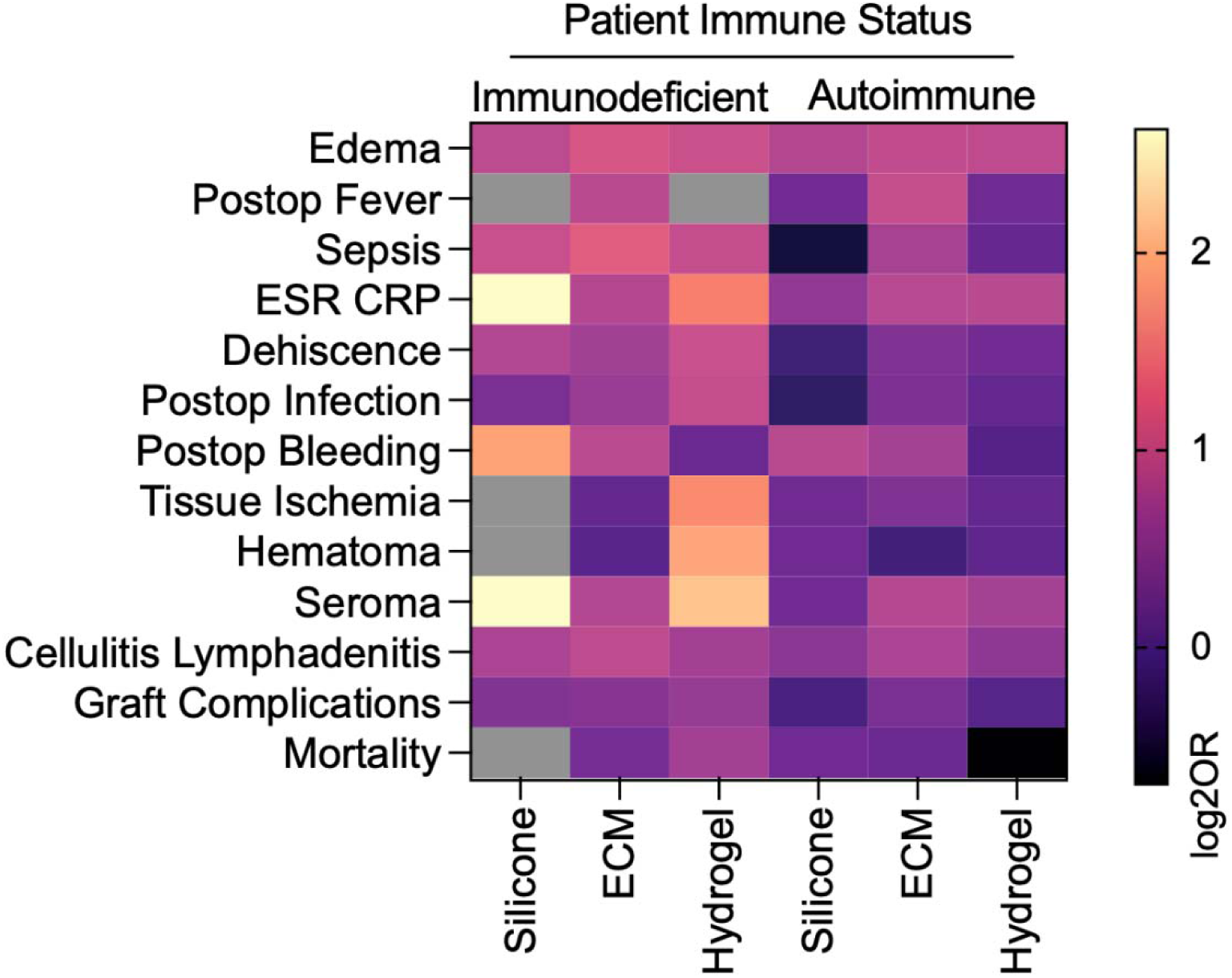
Effect of patient immune status on complications following muscle injury with material implantation. Data are log-transformed odds ratio (log2OR) of immune status compared to immunocompetent controls. Grey boxes represent null event cases in groups tested.

Immunodeficient patients treated with a synthetic hydrogel had higher odds of developing all complications compared to immunocompetent patients, except for post-operative bleeding and mortality (**Figure 2**). Further, immunodeficient patients seemed to experience higher odds of complication compared to autoimmune patients. This was most noticeable for development of sepsis (OR= 1.929, 95%CI, 1.455-2.556) and post-operative infection (OR= 1.933, 95%CI, 1.432-2.61). Autoimmune patients treated with hydrogels, on the other hand, showed minimal differences from immunocompetent controls. We found higher odds of edema (OR= 1.863, 95%CI, 1.562-2.223), ESR/CRP elevation (OR= 1.797, 95%CI, 1.162-2.779), and cellulitis/lymphadenitis (OR= 1.395, 95% CI, 1.232-1.58) only.

Among silicone-treated patients, only the odds of developing local tissue edema were increased for autoimmune patients compared to immunocompetent patients. No other outcomes differed between autoimmune and immunocompetent patients compared to controls or to each other. Immunodeficient patients treated with silicone-based biomaterials had higher odds of developing local tissue edema (OR= 1.857, 95%CI, 1.123-3.069), ESR/CRP elevation (OR= 6.163, 95%CI, 2.545-14.929), post-operative bleeding (OR= 3.612, 95%CI, 1.639-7.691), seroma formation (OR= 6.163, 95% CI, 2.545-14.929) and cellulitis or lymphadenitis (OR= 1.662, 95% CI, 1.2-2.302).

These results suggest that in univariable analysis, immune status is associated with wound healing outcomes following muscle damage predominantly when patients are treated with ECM-based scaffolds. Among autoimmune patients, those who received ECM had the highest odds of complication; complications were similarly low among those receiving silicone or hydrogel.

Among immunodeficient patients, hydrogel treated patients had highest overall odds of complication while silicone-treated patients had lowest complication odds.

### Impact of Type of Muscle Damage on Complications Across Biomaterials and Immune Subgroup

Complications were analyzed across immune subgroups and biomaterials for specific types of muscle damage (**Figure 3**). Silicone-treated patients generally had lowest variability in complications between muscle damage types. Traumatic injury-related muscle damage was paradoxically associated with lower odds of complication in both ECM- and hydrogel-treated patients when compared to other types of muscle damage. In autoimmune ECM- and hydrogel-treated patients, traumatic injury had reduced odds of ESR/CRP elevation, wound dehiscence, and post-operative infection. In ECM-treated immunodeficient patients, injury-related muscle damage was also associated with lower ESR/CRP elevation, wound dehiscence, and post-operative bleeding. Lastly, in immunodeficient patients, injury related muscle damage lowered odds of local tissue edema when treated with silicone, and overall graft complications when treated with hydrogel. On the other hand, traumatic injury increased odds of post-operative fever and overall graft complications in ECM treated autoimmune patients and mortality in ECM treated immunodeficient patients.

**Figure 3.**
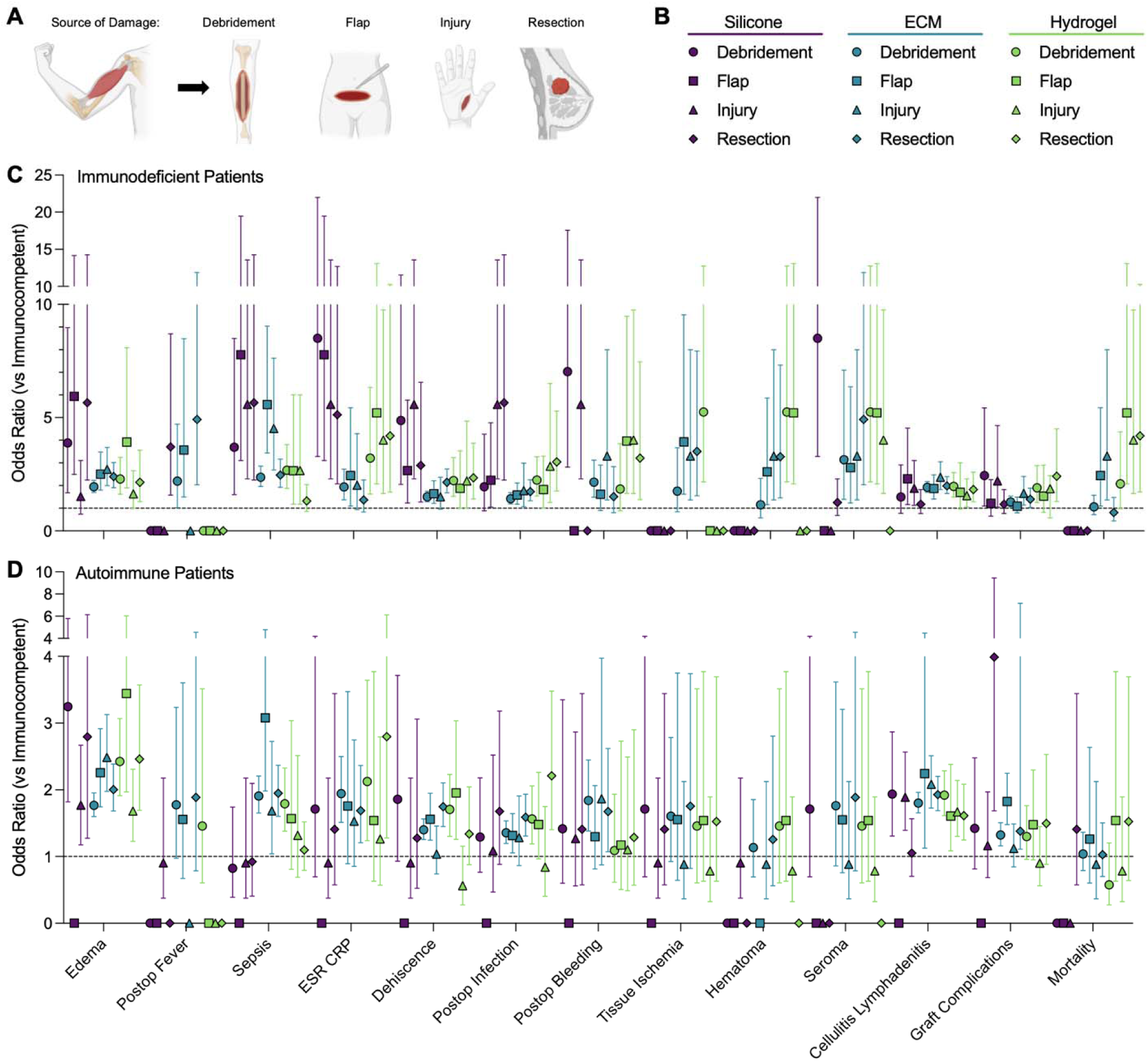
Muscle damage source alters the immune-dependent complications afte material implantation. (A) Subsetting of different muscle damage sources. (B) Key for subgroups analyzed. (C) Odds Ratio (OR) of immunodeficient patients versus immunocompetent. (D) OR of autoimmune patients versus immunocompetent. Data are point estimate ± 95% confidence intervals. Points on x-axis represent null event cases in groups tested. Exact estimates and confidence intervals not shown for n < 10 patients.

Flap harvest and surgical resection related muscle damage types were often associated with increased odds of complication across immune subgroups in patients treated with hydrogel or ECM. In autoimmune patients, flaps and surgical resection were associated with higher odds of post-operative fever, post-operative bleeding, and wound dehiscence in hydrogel- or ECM-treated patients. Among ECM-treated immunodeficient patients, free flap muscle damage conferred higher odds of mortality and graft complications, while hydrogel-treated immunodeficient patients with flap muscle damage had lower odds of graft complications.

In contrast, muscle loss secondary to surgical resection had relatively lower odds of sepsis and post-operative infection, particularly in immunodeficient patients treated with hydrogel. In autoimmune patients, flaps also reduced ESR/CRP elevation and development of cellulitis/lymphadenitis in ECM- and hydrogel-treated patients. Debridement muscle damage showed mixed effects. In autoimmune patients, debridement was associated with increased odds of sepsis and post-operative bleeding when treated with ECM scaffolds and wound dehiscence when treated with hydrogel. Conversely, in immunodeficient patients, debridement was protective in several contexts, with lower odds of infection (silicone), tissue ischemia and hematoma (ECM), and post-operative bleeding and mortality (hydrogel).

Several outcomes consistently showed minimal variation by muscle damage type across biomaterials and immune statuses (edema, sepsis, tissue ischemia, mortality, hematoma, and seroma formation).

### Impact of Immunodeficiency Subtype on Odds of Complication Across Biomaterials in Immunodeficient Patients

Immunodeficiency was further categorized into primary and secondary groups. Primary immunodeficiency included congenital or inherited disorders, including severe combined immunodeficiency (SCID), common variable immunodeficiency, DiGeorge syndrome, Bruton’s agammaglobulinemia, etc. Secondary immunodeficiencies included acquired causes such as HIV, chemotherapy related immunosuppression, and drug-related immunosuppression.

Immunodeficiencies affecting the innate and adaptive immune system were compared (**Figure 4B**). Innate immunodeficiencies included complement disorders, chronic granulomatous disease, leukocyte adhesion defects, etc., while adaptive immunodeficiencies included all T and B cell disorders. Lastly, T cell mediated, and B cell mediated immunodeficiencies were also compared. T cell immunodeficiencies included DiGeorge syndrome thymic aplasia, low CD4+ T cell count associated with HIV disease, Wiskott-Aldrich syndrome and more. B cell immunodeficiencies included Bruton’s agammaglobulinemia, selective IgA deficiency, common variable immunodeficiency, etc. ECM- and hydrogel-treated patients demonstrated more variation based on immune compartment (T cell vs B cell, innate vs adaptive), while silicone-treated patients showed selective differences primarily when comparing primary and secondary immunodeficiencies.

**Figure 4.**
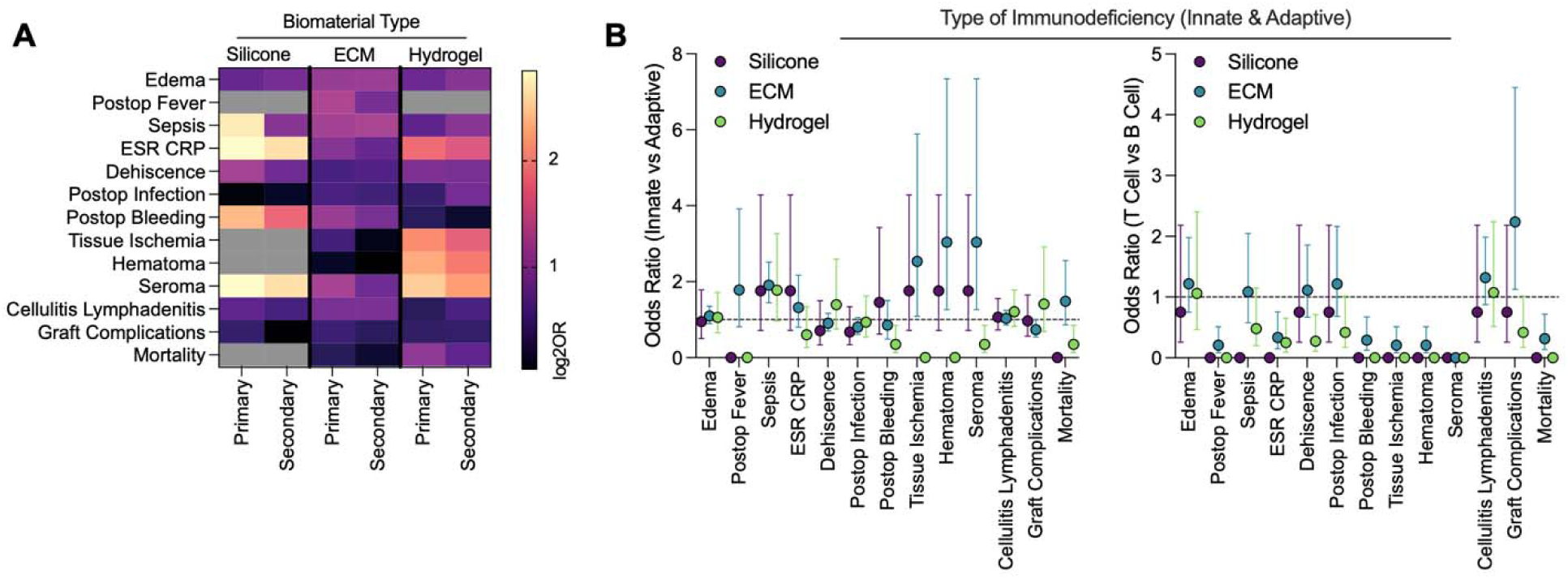
Effect of immunodeficiency type is dependent upon biomaterial type. (A) Primary versus secondary (acquired) immunodeficiencies. Grey boxes represent null event cases in groups tested. (B) Innate and adaptive immunodeficiencies. Data are point estimate ± 95% confidence intervals. Points on x-axis represent null event cases in groups tested. Exact estimates and confidence intervals not shown for n < 10 patients.

Among silicone-treated patients, primary immunodeficiency was most consistently associated with higher odds of complication compared to secondary immunodeficiency for local edema, sepsis, ESR/CRP elevation, and wound dehiscence, with no observed differences by T vs B cell or innate vs adaptive immune classification. Odds of complication in ECM-treated patients demonstrated greater sensitivity to T vs B cell phenotypes, with T cell immunodeficiencies being associated with lower odds of fever, ESR/CRP elevation, dehiscence, bleeding, mortality, tissue ischemia, and hematoma. ECM-treated patients also exhibited higher odds of hematoma, tissue ischemia, and seroma in the setting of innate immune system dysfunction compared to adaptive immune system dysfunction. However, overall graft complication odds, encompassing many causes of graft loss, were higher in T cell immunodeficiencies and adaptive immunodeficiencies. Hydrogel-treated patients showed fewer consistent patterns when comparing immunodeficiency subtypes. Secondary immunodeficiency was associated with higher odds of post-operative infection. T cell dysfunction conferred lower odds of ESR/CRP elevation. Innate immune dysfunction was also linked to lower mortality and seroma risk. Across all biomaterials, complications such as infection, cellulitis/lymphadenitis, and graft complications generally showed minimal variation by immunodeficiency classification, suggesting outcome-specific immune sensitivity.

### Impact of Age on Odds of Complication Across Biomaterials and Immune Phenotypes

Age-related variation in odds of complication differed by biomaterial type and immune subgroup (**Figure 5**). These trends were most pronounced in ECM-treated patients, while hydrogel and silicone groups demonstrated relatively fewer age-related differences. In immunodeficient patients, younger age (18–35) was consistently associated with higher odds of several complications, particularly among those treated with ECM or hydrogel. These included local edema, post-operative fever, ESR/CRP elevation, tissue ischemia, and seroma formation. Among autoimmune patients, increased odds of complication at younger ages were more limited, with elevated risks observed only for fever and tissue ischemia in ECM-treated patients. Notably, age-related variation was largely absent in autoimmune patients treated with hydrogel or silicone. Older age (50-65, and 65+) was associated with increased odds of complications in ECM-treated autoimmune patients, with higher odds of post-operative infection, bleeding, and sepsis, and overall graft complications. These associations were not observed in hydrogel or silicone-treated autoimmune patients, suggesting there may be an age-related risk profile specific to ECM use in the context of autoimmunity.

**Figure 5.**
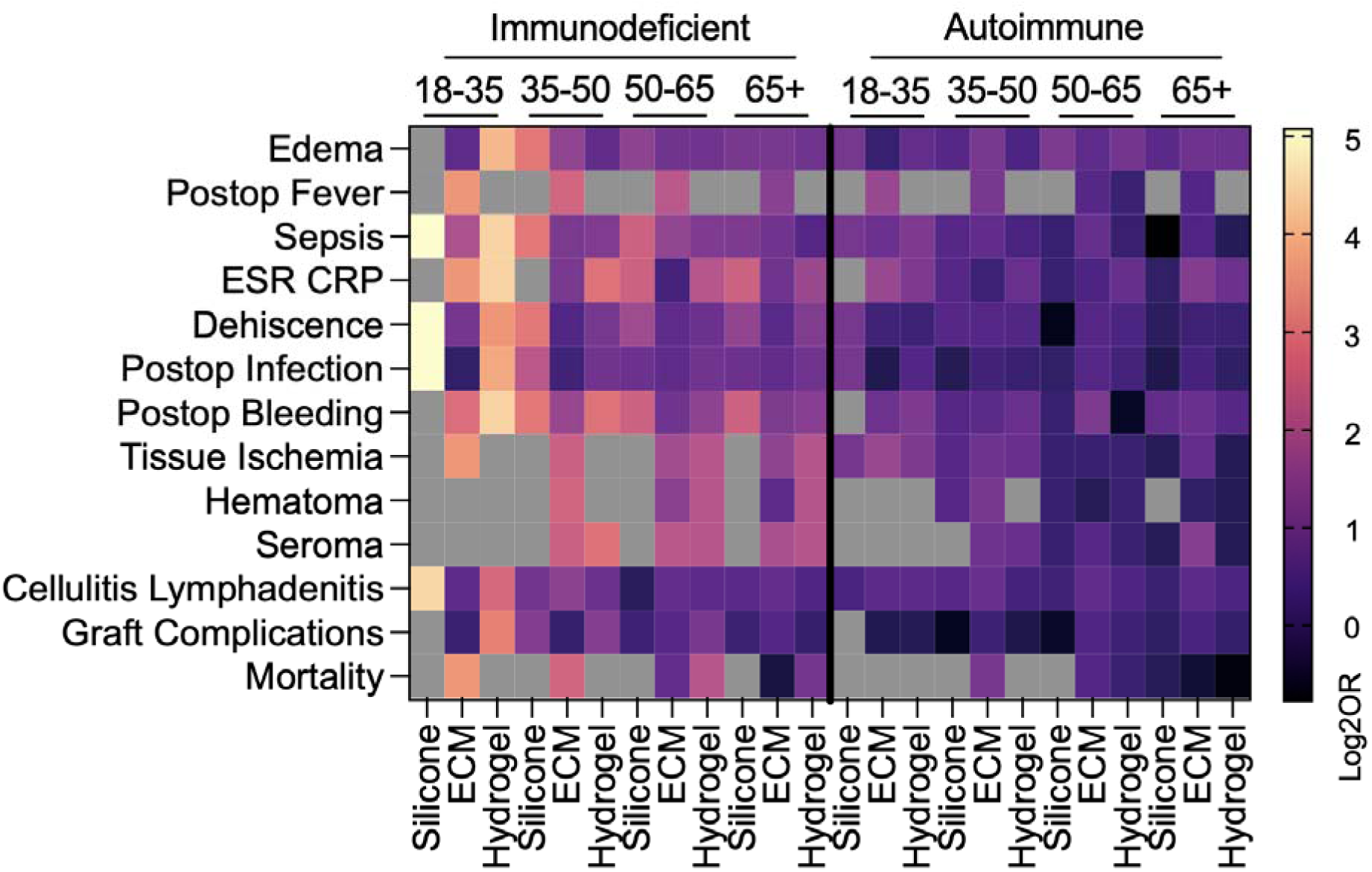
Effects of patient immune status are dependent upon patient age. Data are log transformed point estimate (log2OR) versus immunocompetent. Grey boxes represent null event cases in groups tested.

Some complications exhibited bimodal age-related risk patterns. For instance, ESR/CRP elevation and overall graft complications were more likely to occur at both age extremes (18–35 and ≥65) among ECM-treated patients across immune phenotypes. Lastly, for wound dehiscence, tissue ischemia, and seroma formation, increasing age conferred a protective effect. Wound dehiscence odds decreased with age in silicone-treated patients, and both tissue ischemia and seroma formation decreased with age in immunodeficient patients treated with ECM or hydrogel. No age-related variation was observed in mortality, hematoma formation, or cellulitis/lymphadenitis across any biomaterial type or immune phenotype.

### Impact of Sex on Odds of Complication Across Biomaterials and Immune Phenotypes

Male and female patients were compared for each biomaterial and immune phenotype (**Figure 6**). Sex-related variation in odds of complication varied by complication type, immune status, and biomaterial, with the most notable patterns observed in silicone and hydrogel treated patients. ECM treated patients showed fewer sex-based differences overall. For several complications, male sex was associated with increased odds of adverse outcomes, particularly among autoimmune and immunodeficient patients. Autoimmune male patients had higher odds of local edema, ESR/CRP elevation, and wound dehiscence when treated with hydrogel, higher odds of tissue ischemia when treated with silicone, and higher odds of post-operative bleeding when treated with ECM. In immunodeficient patients, male patients had higher odds of developing sepsis, post-operative bleeding, post-operative infection, ESR/CRP elevation and wound dehiscence in silicone-treated patients. This is likely given the variability in clinical context of silicone material use between males and females. Notably, no sex-based differences were observed in ECM-treated immunodeficient patients across these outcomes. This suggests that among patients with immune dysfunction, sex-related differences play a role in modulating response to biomaterials, specifically hydrogels and silicone-based implants.

**Figure 6.**
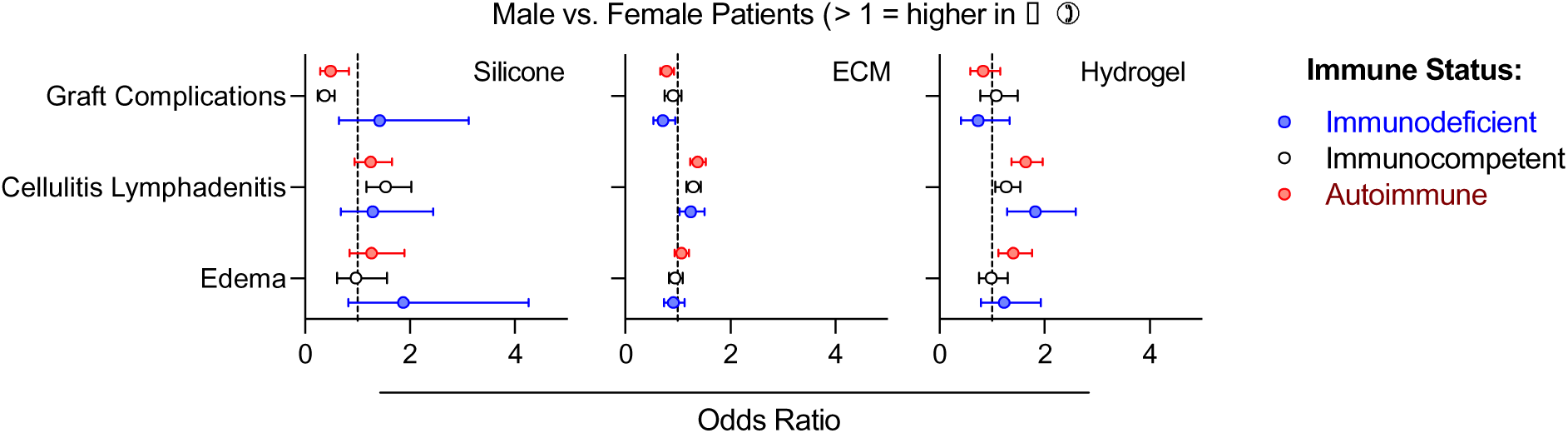
Sex-dependent alterations in complications following muscle injury. Data are odds ratio point estimate ± 95% confidence interval of male versus female patients.

**Figure 7.**
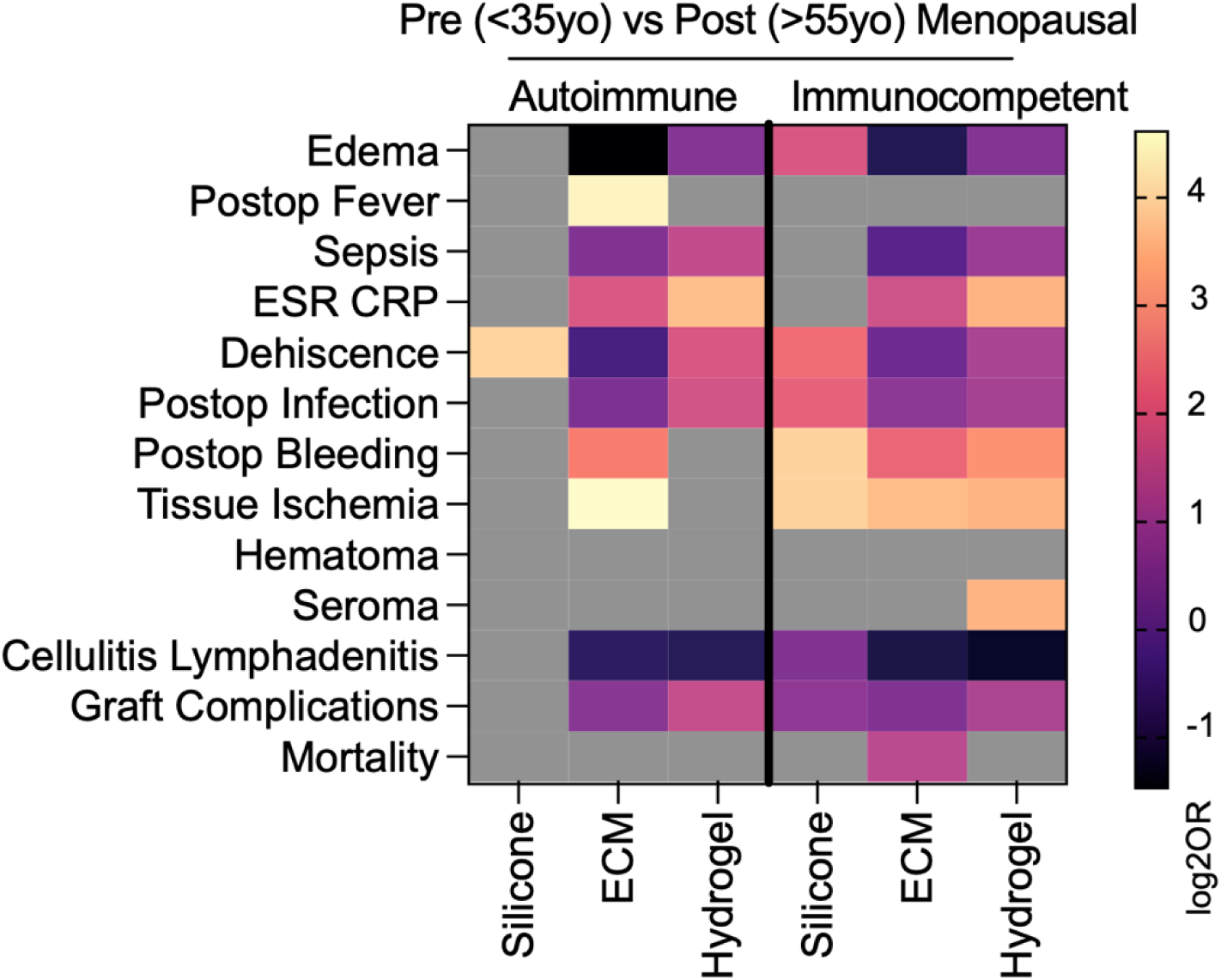
Age related differences of complications in pre-and post-menopausal aged women. Data are log transformed point estimate (log2OR) versus post-menopausal (>55yo). Grey boxes represent null event cases in groups tested.

Among immunocompetent patients, males had lower odds of wound dehiscence with silicone, and higher odds of cellulitis/lymphadenitis with ECM, hydrogel, and silicone. Cellulitis/lymphadenitis consistently showed higher odds in males across immune statuses in both ECM and hydrogel treated groups, and in immunocompetent patients treated with silicone. Conversely, female sex was associated with higher odds of overall graft complications. Males had lower odds of graft complications compared to females in both autoimmune and immunodeficient patients treated with ECM, as well as in autoimmune and immunocompetent patients treated with silicone. No sex-related differences were observed in mortality, hematoma formation, or seroma formation.

### Impact of Age Associated Menopausal Status on Odds of Complication Across Biomaterials in Autoimmune Patients

Menopausal status is a known factor impacting outcomes in autoimmune patients. As such autoimmune patients were split into pre-menopausal (18–35) and post-menopausal (50+) age groups for each biomaterial, excluding ages that may include perimenopause or individual variation during perimenopause ages. Immunocompetent patients were also similarly grouped for comparison. Menopause status impacted complication risk differently across biomaterials and immune phenotypes, with hydrogel and ECM treated patients exhibiting the most consistent postmenopausal improvement. Across nearly all complications, hydrogel treated patients showed higher odds of complication in pre-menopausal patients, though this effect was much more pronounced in immunocompetent patients than autoimmune patients. Pre-menopausal immunocompetent individuals had higher rates of sepsis, ESR/CRP elevation, dehiscence, post-operative infection, post-operative bleeding, tissue ischemia, seroma formation, and graft complications. These findings suggest a potential protective effect of menopause in hydrogel associated wound healing and immune modulation.

ECM treated patients also showed a general trend of higher odds of complication in pre-menopausal patients, especially among the immunocompetent group. These included local edema, ESR/CRP elevation, post-operative infection, post-operative bleeding, tissue ischemia, and mortality. In autoimmune patients, only ESR/CRP, bleeding, and ischemia were elevated in the pre-menopausal group. Interestingly, autoimmune pre-menopausal patients had lower odds of post-operative infection, post-operative bleeding, and tissue ischemia compared to their immunocompetent counterparts across both ECM and hydrogel treatment.

In contrast, silicone treated patients showed fewer menopausal differences, primarily limited to immunocompetent patients. Premenopausal immunocompetent patients exhibited higher rates of local edema, wound dehiscence, post-operative infection, post-operative bleeding, and tissue ischemia, whereas autoimmune patients showed no variation for these complications. Interestingly, cellulitis/lymphadenitis odds were higher in post-menopausal autoimmune patients only, in contrast to other complications. Several outcomes, including post-operative fever, mortality, and hematoma formation, were largely unaffected by menopausal status across all biomaterials and immune groups, suggesting that these complications may be less hormonally modulated.

Given that pre-menopausal autoimmune paradoxically patients exhibited fewer post-operative complications compared to pre-menopausal immunocompetent patients across ECM and hydrogel biomaterial groups, and pre-menopausal patients generally had worse outcomes compared to post-menopausal patients, additional data on medication history in these patient groups was gathered to assess for potential confounders (**Supplemental Table 1**). Non-steroidal anti-inflammatory drugs were prescribed to pre-menopausal patients at greater rates than post-menopausal patients across all biomaterial groups. Further, non-steroidal immunosuppressant medications such as mycophenolate mofetil were also prescribed more to pre-menopausal patients. Conversely, glucocorticoids were prescribed to post-menopausal patients to a greater degree.

## DISCUSSION

In healthy immunocompetent patients, skeletal muscle injuries cause myocyte necrosis leading to neutrophil accumulation followed by macrophage infiltration and satellite cell proliferation(15, 16). Neutrophil proliferation peaks approximately 48 hours after injury and by five days following injury, macrophage polarization transitions from M1 (inflammatory phenotype) to M2 (anti-inflammatory phenotype). Satellite cell proliferation also peaks at this time and regeneration begins. However, this transition can be delayed or impaired in patients with immune imbalances which can lead to aberrant wound healing. In patients with autoimmune diseases, persistent infiltration of inflammatory cells can cause muscle fibrosis and excess fatty deposition. Furthermore, surgical procedures and wounds are known trigger events for autoimmune conditions showing the intricate relationship between tissue damage and autoimmunity(17). In immunocompromised patients, the immune system is unable to sufficiently mediate tissue regeneration and angiogenesis, leading to delayed wound healing and susceptibility to infections.

This study supports these findings, with autoimmune and immunodeficient patients having higher odds of developing nearly all wound healing complications regardless of biomaterial type used in reconstruction in univariable analysis. Autoimmune and immunodeficient statuses did not differ in the degree to which they raised odds of complications, except for infectious complications. While immunodeficiency and immunosuppression are typically considered by surgeons when developing treatment plans, autoimmune disease is not usually considered. Our findings suggest that wound healing following muscle trauma could be similarly impaired in both groups and that any immune-modifying condition should be considered when planning surgeries that require implantation of biomaterials.

One notable difference in complications was found between immune statuses: individuals with autoimmunity were at higher risk of developing post-operative fever and seroma following muscle damage compared to immunocompetent controls, while immunodeficient patients were not. Early post-operative fever is not usually infectious in etiology and can be caused by innate immune system activation and release of pro-inflammatory mediators like interleukin (IL)-1b, IL-6, and TNF-a(15). In autoimmune patients, this response can be exaggerated and can even lead to chronic inflammation, positing a theory for this finding(18). Similarly, seroma formation is mediated by immune dysfunction, with a study in post-mastectomy patients finding that seromas displayed an increase in T-helper cells within the aspirated fluid and also showed elevated Th2 and Th17 responses peripherally(19, 20).

### Biomaterial Type

While there were differences in patients receiving different biomaterial treatments, for example ECM-treated patients had more pre-injury comorbidities than silicone-implanted patients, it seemed that immune status was a larger determinant of receiving ECM. The host response to biologic ECM scaffolds in immunocompetent hosts involves both innate and adaptive immunity but varies based on specific composition(21). A murine model of abdominal wall muscle repair using commercially available native SIS-ECM showed an intense neutrophil and mononuclear cell response with M2 predominance at two weeks following surgery(9). Muscle tissue grafts also became largely well-organized with collagenous connective tissue. However, modifications of grafts with chemical crosslinking led to M1 predominance and increased fibrosis. As such, biologic composition and degradability have an impact on the host immune response to ECM scaffolds. In immunocompromised patients, the foreign body response is minimized, while in autoimmune patients, a significant foreign body response can be seen. A study in mice showed a decrease in M2 polarization with a corresponding increase in adipogenesis in both T cell lacking and autoimmune-prone hosts in response to ECM implantation(8, 11). These findings may explain why those in the autoimmunity subgroup had the highest odds of complication in ECM-treated patients compared to other biomaterials, as ECM components dynamically interact with the immune system. However, clinical studies of immune responses to ECM scaffolds and impacts on wound healing and muscle regeneration are limited often not including individuals with immune imbalance. The present study does not account for alterations in patient comorbidities, suggesting a need for a prospective study to fully evaluate this effect.

Immunodeficient patients, on the other hand, had highest overall odds of complication among those treated with hydrogel and lowest among those treated with silicone, compared to immunocompetent controls. Immune response to hydrogels varies based upon physical properties such as stiffness and topography and chemical properties like molecular presentation and delivery of bioactive molecules(12, 22). Commercially available hydrogel wound fillers can be natural, like alginate or gelatin-based, or synthetic like polyvinyl alcohol or polyethylene glycol-based. Response to these biomaterials in the setting of immunosuppression can be better understood with studies assessing delivery of glucocorticoids within biomaterial implants.

This response, in addition to the clear effect of immunodeficiency on infectious complications, could explain the elevated odds of wound healing complications in immunodeficient patients observed in our cohort. Limited studies have assessed how specific biomaterials could affect this outcome. Biological mechanisms for this phenomenon in individuals treated with hydrogels are unclear given the variability in responses based upon the material composition of the hydrogel. Silicone, on the other hand, may have lower odds of complication in immunodeficient patients compared to other biomaterials given that silicone is known to elicit a strong foreign body response and chronic inflammation in immunocompetent individuals, which is significantly reduced in the setting of immunosuppression (23).

### Muscle Damage Type

Traumatic injury-related muscle damage was paradoxically associated with lower odds of complication in ECM- and hydrogel-treated patients in our cohort when compared to surgical muscle damage. Traumatic injuries typically carry greater risk of post-operative complications due to the nature of their provenance – they are acute, unplanned emergencies that are frequently contaminated by the environment from which they resulted (*e.g.,* dirt, debris, bacteria).^24^ However, in our cohort, injury-associated damage carried lower odds of several complications including ESR/CRP elevation, wound dehiscence, and post-operative infection. This may be explained by aggressive protocols at many hospitals to prevent surgical site infections in trauma populations, including IV antibiotics, delayed wound closure, and inpatient monitoring for severe wounds. Additionally, our surgical muscle damage groups included flap harvest and surgical muscle resection. Surgical resection of muscle is most often done for pathologies like sarcoma to achieve negative margins. Flap harvest is often also done to reconstruct a recipient site where a defect often occurs due to cancer resection or trauma. Given that many surgical modes of muscle damage are done in cases of malignancy, odds of complication may be higher in these groups due to confounders such as adjuvant chemotherapy or radiation and systemic immune dysfunction in the setting of malignancy.

### Mechanism of Immunodeficiency

When comparing the mechanism of immunodeficiency, we found increased odds of complications across primary and secondary immunodeficiencies treated with silicone and hydrogel, compared to immunocompetent controls. Comparing innate deficiencies to adaptive immune deficiencies, there was a trend of higher odds ratios overall with innate immunodeficiencies which aligns with the critical role of innate immune cells like macrophages in wound healing and biomaterials responses. Interestingly, when evaluating T cell versus B cell immunodeficiencies, ECM stood out as having a higher risk of graft complications in T cell immunodeficiencies which is supported by preclinical studies supporting the role of T cells in immune responses to ECM scaffolds.

### Sex Differences

Sex differences in odds of complication found in our study could be due to the immunoregulatory effects of estrogen. Estrogen plays a significant role in modulating immune responses and enhancing wound healing. It promotes anti-inflammatory cytokine production, supports angiogenesis, and facilitates resolution of inflammation (24). These processes are crucial for the successful integration of biomaterials like ECM and hydrogels. Females with higher circulating estrogen levels may suppress immune responses to implanted biomaterials, thereby reducing the risk of inflammatory complications and poor integration when compared to males. Studies in ovariectomized mice, for example show excessive inflammation and impaired wound healing (25). This effect is largely preserved across immune subgroups in our cohort. Furthermore, sex related confounders may also explain these findings. Mechanism of injury, prevalence of behaviors like smoking, and past medical history factors differ in males and females, and the effect of gender may be confounded with these effects. For example, there is a greater prevalence of cigarette smoking in males, and smoking is known to alter outcomes in wound dehiscence (26).

### Age and Menopause Effects

Age is a known factor in modulating immune functioning. Aging is associated with diminished T cell function, altered macrophage activation, and impaired wound healing, which may compromise ECM-mediated tissue remodeling and lead to excessive inflammation or delayed resolution(27, 28). ECM biomaterials, unlike hydrogels and silicone-based implants, rely on active host remodeling and immune cell infiltration, making older patients particularly vulnerable to poor outcomes, especially when immune dysfunction is present(27). This likely accounts for the absence of significant age-related variation in autoimmune patients treated with hydrogels or silicone compared to ECM treated patients. Collectively, these findings highlight the need to consider both patient immune phenotype and age when selecting biomaterials for reconstruction following muscle injury.

In post-menopausal women, the immunosenescence seen with aging can be exacerbated by loss of estrogens(29). However, the clinical impacts of this phenomenon in autoimmune disease are variable. In women with rheumatoid arthritis, menopause can negatively affect the course of disease, while in patients with lupus, menopause leads to improvement in frequency and magnitude of flares. We found increased wound healing complications in pre-menopausal aged women with autoimmune disease compared to this age group in immunocompetent women, which may be due to the variable distribution of specific disease types by age group. In immunocompetent patients, this may be explained by more severe mechanisms of injury such as motor vehicle accidents and other traumatic injuries in younger age groups.

The observation that pre-menopausal aged autoimmune patients treated with ECM and hydrogel biomaterials experience fewer complications than immunocompetent counterparts when directly compared may seem counterintuitive. However, the protective impact of immunosuppressive therapies commonly used in autoimmune disease management may explain these findings. Pre-menopausal women with autoimmune disease were predominantly treated with non-steroidal immunosuppression compared to post-menopausal women. Disease-modifying antirheumatic drugs (DMARDs) dampen both systemic and local immune activity. These therapies may inadvertently provide a protective effect against the foreign body response and inflammation typically associated with biomaterial implantation. In contrast, immunocompetent patients may mount a more robust and prolonged immune response to ECM or hydrogel materials, increasing the likelihood of complications such as dehiscence, seroma, and graft failure.

### Limitations

Due to the retrospective observational nature of this study through a data repository, several limitations must be considered. Firstly, as raw data could not be accessed through the repository due to patient privacy considerations, we were limited in our analyses based on requested comparisons and statistical evaluations. Furthermore, as we were limited in data access, certain comparison weightings could not be made including adjustments for age, sex, and comorbidities. As such, these comparisons are displayed to show potential effects of these variables. Due to the exploratory nature of this work, while corrections for multiple comparisons were completed within variable groups, a more stringent correction would be needed for ongoing studies to evaluate potential false discoveries. Future work will focus on development of specific cohorts in a controlled manner that will allow for in depth statistical characterization based on these initial findings.

## CONCLUSIONS

Through this retrospective observational study utilizing data from EHRs, we supported preclinical data that defines the roles of different immune cell types in wound healing and biomaterial responses. We found that post-operative complications associated with biomaterial treated skeletal muscle injury were highly dependent upon patient immune status when comparing immunocompetent individuals versus those with immunodeficiencies or autoimmune conditions. These increases in post-operative complications were dependent upon patient age, sex, subtype of immune condition, and type of biomaterial implanted.

## AUTHOR CONTRIBUTIONS

SS and KS conceptualized the study; SS analyzed data; SS, AJ, IH, and KS interpreted results; KS provided oversight and funding for the study.

## DATA AVAILABILITY

Electronic health record (EHR) data were analyzed through the TriNetX platform which provides aggregate analyses. All generated odds ratios and estimates provided by TriNetX are available in **Supplemental Data File 1**.

## ACKNOWLEDGEMENTS AND FUNDING

The authors would like to thank Dr. Emily Ricotta for helpful discussions. This work was funded by the intramural research program of the National Institute of Biomedical Imaging and Bioengineering, National Institutes of Health (NIH). <u>Disclaimer</u>: The contributions of the NIH authors are considered Works of the United States Government. The findings and conclusions presented in this paper are those of the authors and do not necessarily reflect the views of the NIH or the U.S. Department of Health and Human Services. Mentioning trade names, commercial products, or organizations does not imply endorsement by the U.S. Government.

